# Dual pandemic of firearm injury and COVID-19 in Central and Southeastern Ohio: An interrupted time series analysis

**DOI:** 10.1101/2025.10.19.25338318

**Authors:** Hannah Williams, Megan Armstrong, Robin Alexander, Jonathan Groner, Bo Lu, Sherri Kovach, Roxanna Giambri, Henry Xiang

## Abstract

**BACKGROUND:** Firearm injuries increased as the United States faced the onset and first two years of the COVID-19 pandemic, a phenomenon some refer to as the “dual pandemic.” Across the country, evidence suggests regional firearm injury rates and related mechanisms behind trends in firearm injury during this time differ, presumably according to local context. This study examines how one regional trauma system fared during the dual pandemic and explores potential explanatory mechanisms of the surge of firearm injuries during the COVID-19 period.

**METHODS:** We used an interrupted time series model to compare quarterly data from 2016-2021 from the COTS (formerly known as the Central Ohio Trauma System) Regional Trauma Registry to examine the proportion of firearm injuries among all trauma cases, case mortality and rates of mistriage among firearm injuries, and the proportion of firearm injuries associated with substance use.

**RESULTS:** Among 3,727 firearm-injured patients, demographic characteristics did not vary with respect to firearm injury, mistriage, mortality, substance use, or alcohol use. Rates of firearm injuries presenting to COTS rose slightly from the beginning of 2020 Q2 (4.54%) through the end of 2020 Q4 (4.71%), and rates continued to grow and remained at a heightened level after the lockdown period. There was no significant change in the growth rate of firearm injuries between 2021 Q2 and 2021 Q3. From 2020 Q4 to 2021 Q2, rates of firearm injury rose 0.99% (CI: 0.05%, 1.9%) from the 2016 average (p<0.05). Considering the entire period from 2016 to 2021, the lockdown period had effects on firearm injury rates that approached significance (p=0.059). There were no significant changes in the rates of undertriage, overtriage, mortality, substance use, or alcohol use. Length of stay did not significantly differ before and during the COVID period.

**CONCLUSION:** This Midwest regional trauma system was affected by the dual pandemic of increased firearm injury during the COVID-19 pandemic. However, the regional trauma system showed resilience, as patient care metrics did not experience significant deficits. Evidence suggests further research into the mechanisms behind significantly increased firearm injury in the region in several months of 2020.

## Introduction

The first two years of the COVID-19 pandemic aligned with a surge in firearm injuries nationwide [1–5], with 2021 holding the greatest amount of firearm injury-related deaths ever recorded [6]. The coinciding public health crises have been referred to as a “dual pandemic” [3]. In 2020 and 2021, trauma systems formed frontlines against both firearm injuries and COVID-19. The COVID-19 pandemic provided unique mechanisms with the possibility to contribute to increased firearm injury, including population mental health challenges, alcohol and other substance use, periods of social unrest, and an influx of firearms into American households.

Increased prevalence of poor mental health offers multiple mechanisms to potentially strain emergency departments (EDs) and trauma centers during the dual pandemic of COVID-19 and firearm injuries. First, increased mental health crises requiring immediate attention would require additional resources from trauma responders. Second, mental health declines may be associated with higher rates of firearm suicide. It is important to note that the pathway from poor mental health to firearm injury is overwhelmingly impacted by firearm suicide and related injuries, not violence directed at others [7, 8], Third, those healthcare workers with an increased risk of COVID-19-related mental health decline might limit the staffing and workload capacities of trauma centers.

The cumulative strains of the pandemic might have led to increased coping through substance use in addition to the existing substance use disorder crises across the country, resulting in associated firearm injuries. Substance use, particularly alcohol use, increases the odds of engaging in firearm-related behaviors, including firearm homicide and self-inflicted firearm injury [9–13]. The sharp increase in alcohol and other substance consumption during the pandemic [14–20] and the significant role of alcohol and other substances in firearm injuries [9–13] suggest the potential for an increase in alcohol-related firearm injuries treated by regional trauma systems in the early years of the pandemic.

Both increases in mental health challenges and substance use occurred during a time of increased firearm purchasing nationwide [21]. Around 3% of all American adults became first-time firearm owners from January 2019 to April 2021 and collectively exposed over 11 million people to household firearms [22], increasing rates of unintentional injury [23], suicide [24], and domestic homicide [25] into affected households [26]. The early months of the pandemic, specifically the lockdown period, saw a dramatic increase in firearm purchasing. From March to mid-July 2020, an estimated 6.5 million adults bought firearms for the first time [27].

The initial wave of COVID-19 across the United States prompted widespread fear and anxiety over the ability of healthcare systems, particularly trauma and EDs, to withstand the threat of critically ill COVID-19 patients en masse. Stay-at-home and physical distancing orders prompted staffing policy shifts [28], potentially limiting the availability and overwhelming the workload capacity of trauma providers [29]. Healthcare workers were burnt out, and hospital resources had to be carefully rationed to avoid depletion [30, 31]. Decreased hospital resource availability during the COVID-19 period was associated with higher mortality for both COVID-19 and non-COVID-19 patients [32–34]. Physicians and political leaders pleaded with their communities to put their firearms down to save beds and hospital resources for COVID-19 patients [35, 36]. Communities were counting on the performance of their local trauma and emergency systems.

Although all-cause mortality rates increased dramatically for hospitals nationwide during the COVID-19 period, trauma centers across different regions in the United States reported varying trauma activation rates [37, 38], injury modalities, and trauma system utilization during the COVID-19 period. Causes of traumatic injury, diagnoses, and procedures were significantly changed by the pandemic [1, 38–40]. Trauma centers nationwide increased use of hospital resources to adjust accordingly to meet the changing demands associated with altered injury patterns [40].

Therefore, this paper examines the response of one regional trauma system and explores some potential mechanisms of the “dual pandemic”. It aims to fill an important role by examining how one regional trauma system, COTS, fared under the weight of the dual pandemic and explores how contextual factors across Central, Southeast, and Southeast Central Ohio may have contributed to the local manifestation of the dual pandemic. Two specific objectives were: (1) to provide an epidemiological description of the dual pandemic of COVID-19 and firearm injury within a regional trauma system, and (2) to identify the effects of simultaneous public health crises on the quality of care delivered by a regional trauma system. We hypothesized that the COTS data would provide evidence of the dual pandemic in the region, more firearm-injured patients would show clinical evidence of substance or alcohol use, and outcomes among those firearm-injured patients would suffer due to increased strain.

## Methods Data Source

COTS (formerly known as the Central Ohio Trauma System) includes two adult Level I trauma centers, one pediatric Level I trauma center, two adult Level II trauma centers, two adult Level III-N and three adult Level III trauma centers, as well as 34 acute care hospitals and 18 free-standing EDs across 37 of Ohio’s 88 counties. Its vast service area encompasses geographically, culturally, and politically diverse regions, including the urban center of Columbus and the surrounding Central Ohio metropolitan area, as well as rural communities in the Appalachian regions of Southeast Ohio. We obtained 2016-2021 regional trauma data from the COTS’s Trauma Registry containing trauma-related pre-hospital, inpatient, and disposition data during the study period. The COTS trauma registry reporting follows the American College of Surgeons’ National Trauma Data Standard with minimal exceptions. Data were accessed for research purposes beginning on January 6, 2023. As the COTS trauma registry contains fully deidentified protected health information, this study was approved by the Nationwide Children’s Hospital Institutional Review Board, which deemed the study ‘No more than minimal risk’ and waived any requirement for informed consent.

## Study Sample

This retrospective, cross-sectional study identified firearm-related injury patients treated within COTS from January 1, 2016, to December 31, 2021, using the International Classification of Diseases, Tenth Revision (ICD-10) code for firearm-related injuries. Any injuries from air guns, gas or spring guns, or pellet guns were excluded. Additionally, all injuries caused by legal intervention were excluded.

We defined the COVID-19 lockdown period as March 16, 2020, to December 20, 2020, to reflect Ohio’s stay-at-home orders and phased reopening and capture a period of increased firearm sales overall and especially among first-time firearm owners. We compared undertriage, overtriage, and case mortality rates of those who had experienced a firearm injury by patient age, sex, race, primary payer, and injury intentions (unintentional, suicide, assault, and other). Additionally, we compared the proportion of firearm-related injuries with evidence of substance use among all firearm-related injuries during the entire study period, as well as drug and alcohol use among those presenting with a firearm-related injury before and after the onset of the COVID-19 pandemic.

## Statistical Analysis

The proportion of firearm-injured patients was calculated for each quarter from 2016 to 2021. We used trauma activation levels (trauma alert 1, 2, 3, or no trauma alert) after hospital arrival, along with the Cribari Matrix,[41] to calculate overtriage and undertriage rates for each quarter.

This study used an interrupted time series model (ITS) [42] to analyze quarterly data on the proportion of firearm-injured patients among all admitted trauma patients, the proportion of assault injuries among all admitted firearm injuries, the undertriage rate, the overtriage rate, and the ED and hospital mortality rate from 2016 to 2021 to investigate changes in trends following the COVID-19 lockdown. This study used quarterly indicators to account for trauma seasonality as well as a level shift indicator from the fourth quarter of 2020 through the second quarter of 2021 to account for the measurable sociocultural impact of the U.S. 2020 Presidential Election and administration change. The undertriage rate, overtriage rate, and ED hospital mortality rate were hypothesized to undergo a temporary level shift from the second quarter of 2020 through the third quarter of 2021. The proportion of firearm injuries was hypothesized to initially increase from the second quarter of 2020 through the second quarter of 2021. From there, the proportion of firearm injuries would remain at a heightened level. The interrupted time series models were conducted in R (version 4), and graphs were created using the ggplot2 package. For quarterly comparisons, Chi-square tests were conducted using SAS version 9.4 (SAS Institute), and a 2-sided *P* < 0.05 was considered statistically significant. To compare firearm injuries associated with substance and alcohol use, the same ITS procedure was followed. This study used a Wilcoxon rank sum test to determine if the mean length of stay differed between the COVID-19 and non-COVID-19 periods.

## Results

A total of 3,727 firearm injury patients, were included. Sex, age group, race, and primary payer did not vary with respect to firearm injury, mistriage, hospital mortality, substance use, or alcohol use rates (Table 1). During the lockdown period, the proportion of firearm injuries slightly increased from the beginning of the second quarter of 2020 (4.54%) through the end of the fourth quarter of 2020 (4.71%). After accounting for seasonality, there was no significant change in the growth rate of the proportion of firearm injuries among all admitted trauma patients between the second quarter of 2021 through the third quarter of 2021. However, from the fourth quarter of 2020 through the second quarter of 2021, during the 2020 US election, the proportion of firearm injuries increased 0.99% (CI: 0.05%, 1.9%) from the 2016 average (p < 0.05). The proportion of firearm injuries among all trauma patients continued to grow and remained at a heightened level after the lockdown period (Figure 1). Considering the entire period from 2016 to 2021, the effect of the COVID-19 lockdown on the proportion of firearm injuries among all trauma patients approached significance (p = 0.0594). There were no significant changes in the undertriage, overtriage, ED hospital mortality rates of firearm-injured patients, the proportion of firearm injuries with clinical evidence of substance use, or the proportion of firearm injuries with clinical evidence of alcohol use from January 2016 to December 2021 (Table 2; Figure 1). Analysis of the Autocorrelation Function (ACF) and the Partial Autocorrelation Function (PACF) series residuals revealed these time series as appropriate analyses of each outcome as a function of the lockdown period. Length of stay did not significantly differ between before and during the COVID period (Table 3).

**Figure 1.**
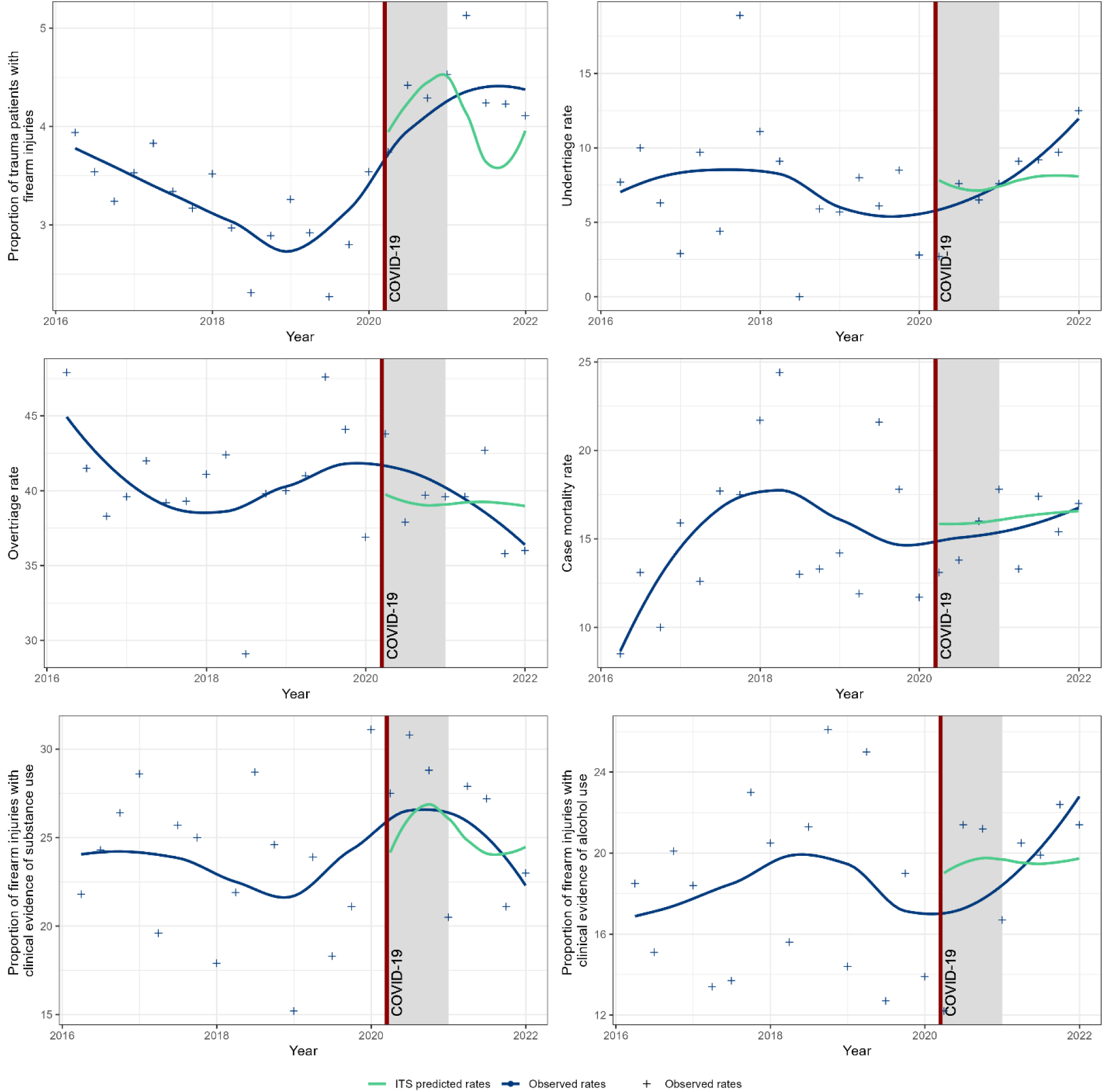
Observed proportion of trauma patients with firearm injuries versus interrupted time series (ITS) predicted rates after the COVID-19 lockdown period in Central, Southeast, and Southeast Central Ohio, 2016-2021.

**Table 1.** Characteristics of firearm injuries, undertriage rates, and mortality during non-COVID and COVID-19 periods in in Central, Southeast, and Southeast Central Ohio, 2016-2021.

|  | Sample (N = 3727) |  |  | Undertriage Rate of Patients with ISS>=10 |  |  | Emergency Department & Hospital Mortality Rate of All Patients |  |  |
| --- | --- | --- | --- | --- | --- | --- | --- | --- | --- |
| Patient Demographics | Non-COVID-19 Period (N, %) | COVID-19 Period (N, %) | P-value | Non-COVID-19 Period (N, %) | COVID-19 Period (N, %) | P-value | Non-COVID-19 Period (N, %) | COVID-19 Period (N, %) | P-value |
| **Sex** |  |  | 0.46 |  |  |  |  |  |  |
| Male | 2628 (85.5%) | 552 (84.4%) |  | 985(5.5%) | 216(3.7%) | 0.29 | 408(15.6%) | 90(16.3%) | 0.64 |
| Female | 445 (14.5%) | 102 (15.6%) |  | 164(6.9%) | 39(12.8%) | 0.41 | 445(13.9%) | 102(13.7%) | 0.96 |
| **Age Group (Yrs)** |  |  | 0.31 |  |  |  |  |  |  |
| 0-17 | 379 (12.3%) | 94 (14.4%) |  | 122(8.2%) | 37(2.7%) | 0.25 | 46(12.1%) | 14(14.9%) | 0.47 |
| 18-64 | 2570 (83.6%) | 533 (81.5%) |  | 993(5.2%) | 205(5.4%) | 0.94 | 398(15.5%) | 82(15.4%) | 0.95 |
| 65+ | 123 (4.0%) | 26 (4.0%) |  | 34(17.7%) | 13(7.7%) | 0.39 | 26(21.1%) | 7(26.9%) | 0.52 |
| Missing | 1 (0.03%) | 1 (0.2%) |  | * | * |  | * | * |  |
| **Race** |  |  | 0.95 |  |  |  |  |  |  |
| White | 1164 (37.9%) | 2435(37.5%) |  | 395(7.1%) | 88(8.0%) | 0.77 | 174 (14.9%) | 35(14.3%) | 0.79 |
| African American | 1764 (57.4%) | 375 (57.3%) |  | 693(5.3%) | 155(3.9%) | 0.45 | 261(14.8%) | 61(16.3%) | 0.47 |
| Other | 69 (2.3%) | 17 (2.6%) |  | 21 (4.8%) | 6 (0.0%) | 0.58 | 11(15.9%) | 3(17.7%) | 0.86 |
| Missing | 76 (2.5%) | 17(2.6%) |  | * | * |  | 24(31.6%) | 5(29.4%) | 0.86 |
| **Primary Payer** |  |  | 0.89 |  |  |  |  |  |  |
| Government* | 1866 (60.7%) | 404 (61.8%) |  | 656(6.7%) | 169(5.3%) | 0.51 | 213(11.4%) | 54(13.4%) | 0.27 |
| Private insurance | 476 (15.5%) | 105 (16.1%) |  | 165(6.1%) | 33(6.1%) | 1.00 | 65(13.7%) | 12(11.4%) | 0.54 |
| Self-pay | 663 (21.5%) | 133 (20.3%) |  | 303(4.3%) | 50(2.0%) | 0.44 | 181(27.3%) | 38(28.6%) | 0.77 |
| Other | 43(1.4%) | 7(1.1%) |  | 14(0.0%) | 2(0.0%) | * | 3(7.0%) | 0(0.0%) | 0.47 |
| Missing | 27(0.9%) | 5(0.8%) |  | 11(9.1%) | 1 (100%) | * | 8(29.6%) | 0(0.0%) | 0.15 |
| **Injury intent** |  |  | 0.73 |  |  |  |  |  |  |
| Unintentional injuries | 553 (18.0%) | 127 (19.4%) |  | 82(13.4%) | 21(4.8%) | 0.27 | 23(4.2%) | 3(2.4%) | 0.34 |
| Intentional self-harm | 236 (7.7%) | 45 (6.9%) |  | 168(7.1%) | 29(6.9%) | 0.96 | 121(51.3%) | 19(42.2%) | 0.27 |
| Assaults and homicide attempt | 2099 (68.3%) | 446 (68.2%) |  | 834(5.2%) | 193(4.7%) | 0.78 | 302(14.4%) | 77(17.3%) | 0.12 |
| Other (undetermined) | 185 (6.0%) | 36 (5.5%) |  | 65(3.1%) | 12(8.3%) | 0.39 | 24(13.0%) | 5(13.9%) | 0.88 |

**Table 2.** Interrupted time series-predicted rates of change after the COVID-19 lockdown period in Central, Southeast, and Southeast Central Ohio, 2016-2021.

| Model | Estimated effect | Standard Error | p-value ($\alpha = 0.05$) |
| --- | --- | --- | --- |
| Proportion of firearm injuries out of all trauma cases | 0.04339 | 0.02157 | 0.0594 |
| Undertriage rate | -0.03614 | 0.1384 | 0.7965 |
| Overtriage rate | -0.02351 | 0.13701 | 0.865 |
| Case mortality | -0.00656 | 0.13454 | 0.962 |
| Proportion of firearm injuries with clinical evidence of substance use | 0.12326 | 0.15075 | 0.423 |
| Proportion of firearm injuries with clinical evidence of alcohol use | 0.02500 | 0.1416 | 0.862 |

**Table 3.** Length of stay among those with firearm injuries during non-COVID and COVID-19 periods in Central, Southeast, and Southeast Central Ohio, 2016-2021.

|  | N | Mean | Minimum | Maximum | P-value from Wilcoxon Two-Sample Test |
| --- | --- | --- | --- | --- | --- |
| Non-COVID-19 period | 3073 | 4.7 | 1 | 96 | 0.25 |
| COVID-19 period | 654 | 4.3 | 1 | 73 |  |

## Discussion

As evidenced by this study, Central, Southeast, and Southeast Central Ohio experienced a rise in firearm injuries during the COVID-19 that approached significance. Characteristics of those who suffered from firearm injuries, however, did not change during the COVID-19 crisis; Sex, age, race, primary payer, and injury intent had no significant effect on undertriage rates or case mortality. Of all the outcomes analyzed by ITS, only the proportion of firearm injuries increased, albeit just outside of an α of 0.05. This shows that the region served by COTS was not spared from the national phenomenon. In fact, the proportion of those experiencing a firearm injury among all admitted trauma patients reached its highest rates and showed the most significant growth during the months surrounding the U.S. Presidential Election and administration change, hinting at the complicated relationship between firearm injuries and social unrest that should be explored in future studies. While rates of firearm injury rose, undertriage, overtriage, and case mortality remain unchanged by exposure to the COVID-19 lockdown period.

This study’s findings show the resilience of COTS under the strain of the dual pandemic and all of its systematic stressors. Such resilience might be attributed to successful professional guidance such as the American College of Surgeons Committee on Trauma’s “Maintaining Trauma Center Access and Care during the COVID-19 Pandemic: Guidance Document for Trauma Medical Directors” [43]. Much of the observed resilience is owed to the providers and professionals who kept COTS facilities operating during prolonged crises.

In addition to the impact of the dual pandemic on care delivery and quality among firearm-injured patients, this study explored potential mechanisms of increased firearm injury during the COVID-19 pandemic in Central, Southeast, and Southeast Central Ohio. ITS results show two important findings. First, heightened substance and alcohol use during the pandemic did not seem to have a significant effect on cases of firearm injury within this population. The proportion of those with clinical evidence of any substance use did not significantly change, and neither did the proportion of those with clinical evidence of alcohol use. This contradicts evidence of potential mechanisms introduced in this paper [9–13] that highlights the relationships between substance and alcohol use with firearm injuries and increased substance use as coping during the COVID-19 lockdown. These findings are particularly surprising because sharp increases in overdose mortality during the first seven months of the pandemic in Ohio [44] suggest a rapid uptick of substance use in the state following the declaration of the national emergency. Second, the most significant spike in firearm injuries is found near the end of 2020 and the beginning of 2021. This time can be categorized as the period surrounding the 2020 U.S. Presidential Election and 2021 administration change. Further research should investigate how sociopolitical uncertainty may have influenced firearm injury rates.

### Limitations

Although regional data is a key factor of this paper, it prevents us from generalizing our findings to other populations. Additionally, retrospective data and a lack of contextual evidence prevent us from making causal claims regarding the dual pandemic in Ohio, substance use among firearm injuries, and sociopolitical culture surrounding the 2020 election and 2021 administration change. Registry data is subject to variability, considering the large area and variety of institutions serviced by COTS.

## Conclusions

This study analyzed the dual pandemic of COVID-19 and firearm injury at the regional level. Results show that while COTS did face heightened firearm injuries during the COVID-19 pandemic, their performance did not falter. This paper explored how the dual pandemic manifested in this regional trauma system and revealed that Central, Southeast, and Southeast Central Ohio faced increased firearm injury in the period surrounding the 2020 U.S. Presidential Election and 2021 administration change. Further research should identify how and why COTS showed resilience while faced with the dual pandemic and compare its performance to other regional trauma systems. It is necessary to analyze the particular factors contributing to resilience across COTS membership institutions to better tell the story of this remarkable time in history, improve trauma care, and enhance disaster preparedness among regional trauma systems.

## Data Availability

The data that support the findings of this study are available from COTS but restrictions apply to the availability of these data, which were used under license for the current study, and so are not publicly available. COTS (formerly known as the Central Ohio Trauma System) data-sharing policies require persons who are interested in data to reach out to COTS directly.

## Declarations

## Acknowledgements

The authors would like to thank our partners at COTS and the many who work across its service region to care for Ohioans on their hardest days. We would also like to acknowledge Dr. Biche Osong for assisting with the interrupted time series analyses.

## Ethics Approval and Consent to Participate

The Institutional Review Board at Nationwide Children’s Hospital (IRB00000568) approved this study on June 6, 2022, and deemed the study ‘No more than minimal risk’ Human Subjects’ Research.

## Funding

Efforts by Xiang, Armstrong, Alexander, Groner, and Lu in this project were supported in part by a research grant from the National Institute of Health/Eunice Kennedy Shriver National Institute of Child Health and Human Development (R01 HD107280-01A1, Xiang-PI).

The Eunice Kennedy Shriver National Institute of Child Health and Human Development had no role in the design and conduct of the study; collection, management, analysis, and interpretation of the data; preparation, review, or approval of the manuscript; and the decision to submit the manuscript for publication. The contents do not represent the views of the Eunice Kennedy Shriver National Institute of Child Health and Human Development.

## Author Contributions

Dr. Henry Xiang had full access to all of the data in the study and takes responsibility for the integrity of the data and the accuracy of the data analysis.

Concept and design: Xiang, Armstrong, Alexander, Williams

Acquisition, analysis, or interpretation of data: All authors.

Drafting of the manuscript: Williams, Xiang, Armstrong.

Critical revision of the manuscript for important intellectual content: All authors.

Statistical analysis: Xiang, Alexander, Williams.

Obtained funding: Xiang, Groner, Lu.

Administrative, technical, or material support: Armstrong, Kovach, Giambri.

Supervision: Xiang.

